# Balancing Stability and Recovery: Impact of Spinal Fusion Length on Surgical Site Infections and Functional Outcomes in Traumatic Thoracic and Thoracolumbar Spinal Cord Injuries

**DOI:** 10.1101/2024.12.20.24318149

**Authors:** Iris Leister, Moritz Katzensteiner, Christof Wutte, Orpheus Mach, Doris Maier, Matthias Vogel

**Author notes:** Corresponding author: Iris Leister, PhD Spinal Cord Injury Center BG Trauma Center Murnau Professor-Kuentscher-Straße 8 82418 Murnau am Staffelsee, Germany /48-3824.

## Abstract

**Background:** Spinal surgeries after traumatic thoracic or thoracolumbar spinal cord injuries (SCI) often involve extensive instrumentation, leading to an increased risk of surgical site infections (SSIs) and complications. This study investigates whether the number of spinal levels fused impacts functional recovery and the frequency of SSIs in individuals with vertebral fractures in the thoracic and/or lumbar spine and SCI.

**Methods:** In this retrospective analysis of 282 patients with traumatic thoracic or thoracolumbar SCI, data across two periods (2005–2009 vs. 2018–2022) were compared, including variables such as demographics, setting of the initial surgery (specialized SCI center or general trauma center), SCI severity, number of levels fused, fracture classification, the need for revision surgery, the presence of polytrauma and/or traumatic brain injury (TBI), and the occurrence of SSIs during hospitalization. Functional independence was assessed using the Spinal Cord Independence Measure (SCIM III). Unbiased recursive partitioning (URP-CTREE) was used to identify predictors of SSIs and functional outcomes.

**Results:** Patients undergoing longer fusions had a significantly higher rate of SSIs (median fusion length: 5 vs. 3 levels, p = 0.02). Revision surgery and age over 57 were identified as key predictors of SSIs. Shorter fusions (two spinal segments or less) were associated with better functional recovery, particularly in more severe SCI (AIS A, B, or C).

**Conclusion:** Longer fusion lengths increase the risk of SSIs and are linked to poorer functional outcomes in patients with thoracic and thoracolumbar SCI. Minimizing fusion length might optimize surgical outcomes and reduce complications. Future research should refine surgical strategies to balance stability with functional recovery.

## Introduction

Spinal surgeries after vertebral fractures often require invasive approaches, implantation of significant amounts of instrumentation, and long operating times.^1^ In individuals who sustained traumatic spinal cord injury (SCI), an early surgical treatment including spinal cord decompression, fracture reduction, and spinal column stabilization is recommended.^2–4^

Surgical site infections (SSI) pose a serious post-operative complication in spine surgery.^5^ Among other risk factors, the presence of SCI itself is a significant risk factor for SSIs.^2,6,7^ Non-modifiable risk factors include female sex ^2,7^ and the patients’ age with the lowest risk in teenagers, increased risk in young adults, and the highest in those over 60.^5,8^ Partially modifiable, patient-associated pre-operative risk factors are obesity and diabetes, smoking, and poor nutritional status.^6,9–16^ These factors further complicate clinical decision-making in individuals with acute injury requiring immediate treatment.

The surgical approach and the number of levels fused have been identified as treatment-related risk factors of SSI.^2,7^ Surgeries with extensive use of stabilizing instruments are associated with an increased risk for the development of SSIs, ranging from 2.5% to 11.2%.^5,17^ Also, a delay between injury onset and surgical intervention, longer surgery duration, and prior spinal surgery were found to increase the risk of SSIs.^5,6,17–20^ This is particularly problematic for individuals who require revision surgery.^21^ The surgical invasiveness index (SSI II), used to objectify the overall invasiveness of surgical procedures, was found to be the most reliable predictor for SSIs, even after adjusting for confounders like age and comorbidities.^6,22^ Additionally, both posterior ^5^, and combined anterior/posterior spinal approaches ^1,6^ increase the risk factors of SSIs.

The potential sequelae of SSIs include pseudarthrosis, poor neurologic and general outcome, sepsis, and death.^21,23–26^ SSIs consequently deteriorate the individual patients’ medical condition thereby limiting their potential for recovery after traumatic SCI.^1,5^ Additionally, post-operative wound infections and hospital-acquired pneumonia are associated with poor neurological outcomes, worse motor recovery, and greater likelihood of physical dependence in individuals with SCI.^27,28^

The aim of this study was therefore to investigate if the number of levels fused is associated with 1) functional recovery, and 2) the frequency of SSIs in individuals who sustained traumatic fractures of the thoracic and/or lumbar spine and SCI.

## Materials and Methods

We retrospectively analyzed data from individuals with traumatic thoracic or thoracolumbar SCI who were treated in the BG Trauma Center Murnau.

Standards for surgical treatment have changed over time, now recommending short segment instrumentation and favoring minimally invasive techniques for thoracolumbar fractures to minimize surgical invasiveness and reduce complications.^29^ Additionally, in the 2010 update, the Diagnosis-Related Groups (DRG) system introduced new classifications and refinements for spine surgeries to differentiate reimbursement based on the amount and type of instrumentation used. Therefore, we compared two different time periods in our study: 2005-2009 versus 2018-2022.

In total, we identified 353 records of patients who sustained fractures in the thoracic and/or lumbar spine. We excluded data of 27 patients because they did not provide consent to use their medical data for research purposes. An additional 40 cases had to be removed for the following reasons: concomitant neurological diseases, no surgical treatment, shot gun wounds, not enough neurological assessments to quantify changes between admission and discharge, concomitant cancer diagnosis & treatment, death during hospitalization. The total sample size therefore consists of 282 cases.

The following variables were obtained from the EMSCI records: demographic variables (sex, age at injury), initial severity of SCI according to the American Spinal Injury Association (ASIA) Impairment Scale (AIS), and functional independence according to the Spinal Cord Independence Measure (SCIM III).

The following parameters were extracted from computed tomography (CT) images: fracture classification according to the AOSpine subaxial and thoracolumbar spine injury classification system ^30^ and the number of spinal segments fused. In patients with multiple fractures (n=19), we assigned the worst AO classification if all fractures where within the spinal fusion area (n=5). If a case presented with an additional spinal fusion outside of the segments fused, we assigned “multiple” to the variable of fracture classification (n=14). In 44 cases of patients who underwent spine surgery at another hospital, we had no access to pre-operative CT scans and were therefore unable to classify the fractures.

Additionally, we curated the following variables from our clinical information system: if the initial surgical treatment has been performed in our center, or a patient has been transferred for SCI care and received surgical treatment in another hospital or trauma center; time between injury and transfer to a specialized SCI center; if a patient required revision surgery, sustained polytrauma and/or an additional traumatic brain injury (TBI), or presented with a SSI during hospitalization.

### Statistical analysis

All statistical analyses and figures were completed in R (R Core Team, 2023, version 4.2.3). Descriptive statistics for continuous variables and frequency counts for categorical variables were calculated. To test the association between categorical and continuous variables χ², Fisher’s exact, Student’s t-, and Wilcoxon-Mann-Whitney-Test were applied as appropriate. Normality of distribution was determined using Shapiro–Wilk test.

In order to evaluate if parameters related to the fracture type and treatment of spine fractures, in addition to demographic factors (age, sex) and the initial severity of SCI, are predictive of functional outcome, as well as SSIs in individuals with thoracic and thoracolumbar SCI, we used unbiased recursive partitioning by conditional inference (URP-CTREE).^31–33^

We included the following variables in the URP model aiming to predict the occurrence of an SSI (yes/no): age, sex, AIS at admission, vertebral fracture level (thoracic T2-T10 vs. thoracolumbar T11-L5), number of levels fused, polytrauma and/or additional TBI (yes/no), revision surgery (yes/no), and the AO fracture classification. For model building, 44 cases where fracture classification was not assessed, and 14 with multiple fractures were removed from the dataset resulting in an n = 224 patients for the URP with the endpoint of SSI.

For the endpoint SCIM at discharge, we included the following variables in the URP: AIS at admission, vertebral fracture level (thoracic T2-T10 vs. thoracolumbar T11-L5), number of levels fused, polytrauma and/or additional TBI (yes/no), revision surgery (yes/no), and SSI (yes/no). In this model, the fracture classification was not included as no differences were observed between the observation periods or the treating centers, resulting in a sample size of 282 patients for this analysis.

## Results

Patient characteristics and demographics are depicted in table 1 comparing the two time periods (2005-2009 vs. 2018-2022). Significantly more patients sustained polytrauma and/or additional TBI in the observation period between 2018 and 2022, compared to the earlier period between 2005 and 2009 (24.5% vs. 11.1%; Fisher’s exact test p = 0.005). Also, significantly more patients required revision surgery (25.9% in 2018-2022 vs. 9.6% in 2005-2009; Fisher’s exact test p < 0.001), and the number of levels fused was significantly higher in the later period (median 4 vs. 3 spinal levels, Wilcoxon-Mann-Whitney-Test p < 0.001). Sex, age, the initial severity of SCI measured by the AIS, the fracture classification according to AO, the vertebral fracture level (thoracic vs. thoracolumbar), time from SCI to admission to a specialized SCI center, SCIM at discharge, as well as SCIM change during the hospitalization were not different between the two observation periods. Interestingly, SSIs increased from 1.5% to 6.1% (not statistically significant; Fisher’s exact test p=0.063). In total 11 out of 282 (3.9%) patients sustained an SSI (table 1).

**Table 1:** Patient characteristics.

|  | 2005 - 2009<br>(N=135) | 2018 - 2022<br>(N=147) | Overall<br>(N=282) | P-Value |
| --- | --- | --- | --- | --- |
| Sex |  |  |  |  |
| male | 115 (85.2%) | 121 (82.3%) | 236 (83.7%) | 0.524 |
| female | 20 (14.8%) | 26 (17.7%) | 46 (16.3%) |  |
| Age (years) |  |  |  |  |
| Mean (SD) | 41.1 (16.2) | 39.8 (17.0) | 40.4 (16.6) | 0.396 |
| Median [Min, Max] | 41.0 [13.0, 80.0] | 37.0 [16.0, 81.0] | 39.0 [13.0, 81.0] |  |
| AIS at admission |  |  |  |  |
| A | 82 (60.7%) | 77 (52.4%) | 159 (56.4%) | 0.430 |
| B | 10 (7.4%) | 18 (12.2%) | 28 (9.9%) |  |
| C | 14 (10.4%) | 18 (12.2%) | 32 (11.3%) |  |
| D | 29 (21.5%) | 34 (23.1%) | 63 (22.3%) |  |
| AO fracture classification |  |  |  |  |
| A | 22 (16.3%) | 25 (17.0%) | 47 (16.7%) | 0.155 |
| B | 12 (8.9%) | 17 (11.6%) | 29 (10.3%) |  |
| C | 72 (53.3%) | 76 (51.7%) | 148 (52.5%) |  |
| multiple | 3 (2.2%) | 11 (7.5%) | 14 (5.0%) |  |
| not assessed | 26 (19.3%) | 18 (12.2%) | 44 (15.6%) |  |
| Vertebral fracture level |  |  |  |  |
| thoracic T2-T10 | 55 (40.7%) | 59 (40.1%) | 114 (40.4%) | 1.000 |
| thoracolumbar T11-L5 | 80 (59.3%) | 88 (59.9%) | 168 (59.6%) |  |
| Polytrauma and/or TBI |  |  |  |  |
| no | 120 (88.9%) | 111 (75.5%) | 231 (81.9%) | 0.005 |
| yes | 15 (11.1%) | 36 (24.5%) | 51 (18.1%) |  |
| Surgical site infection |  |  |  |  |
| no | 133 (98.5%) | 138 (93.9%) | 271 (96.1%) | 0.063 |
| yes | 2 (1.5%) | 9 (6.1%) | 11 (3.9%) |  |
| Revision surgery |  |  |  |  |
| no | 122 (90.4%) | 109 (74.1%) | 231 (81.9%) | < 0.001 |
| yes | 13 (9.6%) | 38 (25.9%) | 51 (18.1%) |  |
| number of levels fused |  |  |  |  |
| Mean (SD) | 3.28 (1.48) | 3.97 (1.84) | 3.64 (1.71) | 0.001 |
| Median [Min, Max] | 3.00 [1.00, 9.00] | 4.00 [1.00, 11.0] | 4.00 [1.00, 11.0] |  |

|  | 2005 - 2009 | 2018 - 2022 | Overall | P-Value |
| --- | --- | --- | --- | --- |
| time SCI - admission in SCI center (days) |  |  |  |  |
| Mean (SD) | 9.84 (11.4) | 10.6 (12.8) | 10.2 (12.1) | 0.630 |
| Median [Min, Max] | 6.00 [0, 55.0] | 7.00 [0, 77.0] | 6.00 [0, 77.0] |  |
| SCIM at discharge |  |  |  |  |
| Mean (SD) | 68.2 (20.7) | 69.2 (20.7) | 68.7 (20.7) | 0.840 |
| Median [Min, Max] | 71.0 [6.00, 100] | 69.0 [9.00, 100] | 71.0 [6.00, 100] |  |
| change in SCIM between admission and discharge |  |  |  |  |
| Mean (SD) | 41.4 (20.3) | 42.0 (19.0) | 41.7 (19.6) | 0.801 |
| Median [Min, Max] | 46.0 [-1.00, 96.0] | 42.0 [0, 95.0] | 43.0 [-1.00, 96.0] |  |

### Fusion length

The number of spinal segments fused was found to have a significant association with the occurrence of SSIs. Patients who developed an SSI had a significantly longer median fusion length compared to those who did not develop an SSI (median (IQR) = 5(1) vs. 3(3); Wilcoxon-Mann-Whitney-Test W = 893, p = 0.02; Figure 1a). Also, the fusion length was significantly longer in patients with thoracic fractures (T2-T10, n=114) compared to those with thoracolumbar fractures (T11-L5, n=168), with a median (IQR) of 5(1) vs. 2(2) spinal levels, respectively (Wilcoxon-Mann-Whitney test: W = 15465, p < 0.001).

**Figure 1:**
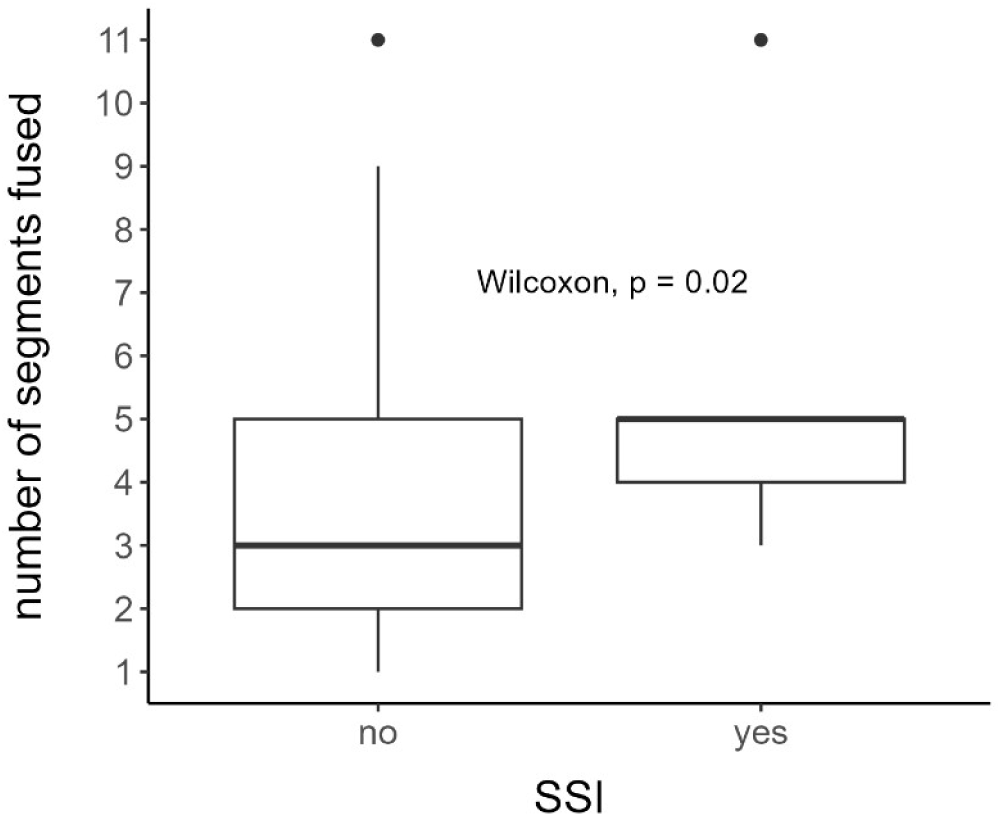
number of fused segments: group comparison SSI vs. no SSI

#### Fusion length over time between the treating centers

While the median fusion length increased by one segment in the overall cohort between the two observation periods (4 vs. 3 spinal levels), the number of levels fused was not significantly different in individuals who underwent their initial spine surgery in a specialized SCI center (n=93, p > 0.05). However, for those who underwent surgery in another hospital and were later transferred to the SCI center (n=189), the fusion length was significantly longer in the period 2018-2022 compared to 2005-2009 (median (IQR): 4(2) vs. 3(2); Wilcoxon-Mann-Whitney Test W = 3273, p = 0.002). The fracture classification, as well as the vertebral fracture level (thoracic vs. thoracolumbar) did not differ between the treating centers, neither overall nor within the observation periods, indicating that the severity and level of the spine fracture did not account for the observed differences in fusion length.

Generally, when comparing the group who received their initial surgery in a specialized SCI center with those who underwent spine surgery in another hospital, the fusion length was significantly longer in the latter (all 282 cases in both observation periods: median (IQR): 4(3) vs. 2(3); Wilcoxon-Mann-Whitney Test W = 10474, p = 0.007). There was no difference in the number of segments fused between the treating centers in the first observation period (2005-2009; p > 0.05). Although, in the more recent period (2018-2022), fusion length was significantly longer in those who underwent fusion in another hospital (median (IQR): 4(2) vs. 3(2); Wilcoxon-Mann-Whitney Test W = 2699, p = 0.02).

### Outcome prediction

#### SSI

The URP for the endpoint of SSIs yielded two cohort splitting nodes: revision surgery and age. Individuals who required revision surgery had a significantly higher chance of obtaining an SSI. Within the group who required revision surgery, individuals above the age of 57 years, also had a significantly higher risk of SSIs (figure 2). SSIs and revision surgery are significantly associated, but the bidirectional relationship makes it hard to determine causality—whether revision surgery caused the SSI or vice versa. In our cohort, significantly more patients initially treated at other hospitals required revision surgery compared to those who had their first surgery in the SCI center (Fisher’s exact test p < 0.001).

**Figure 2:**
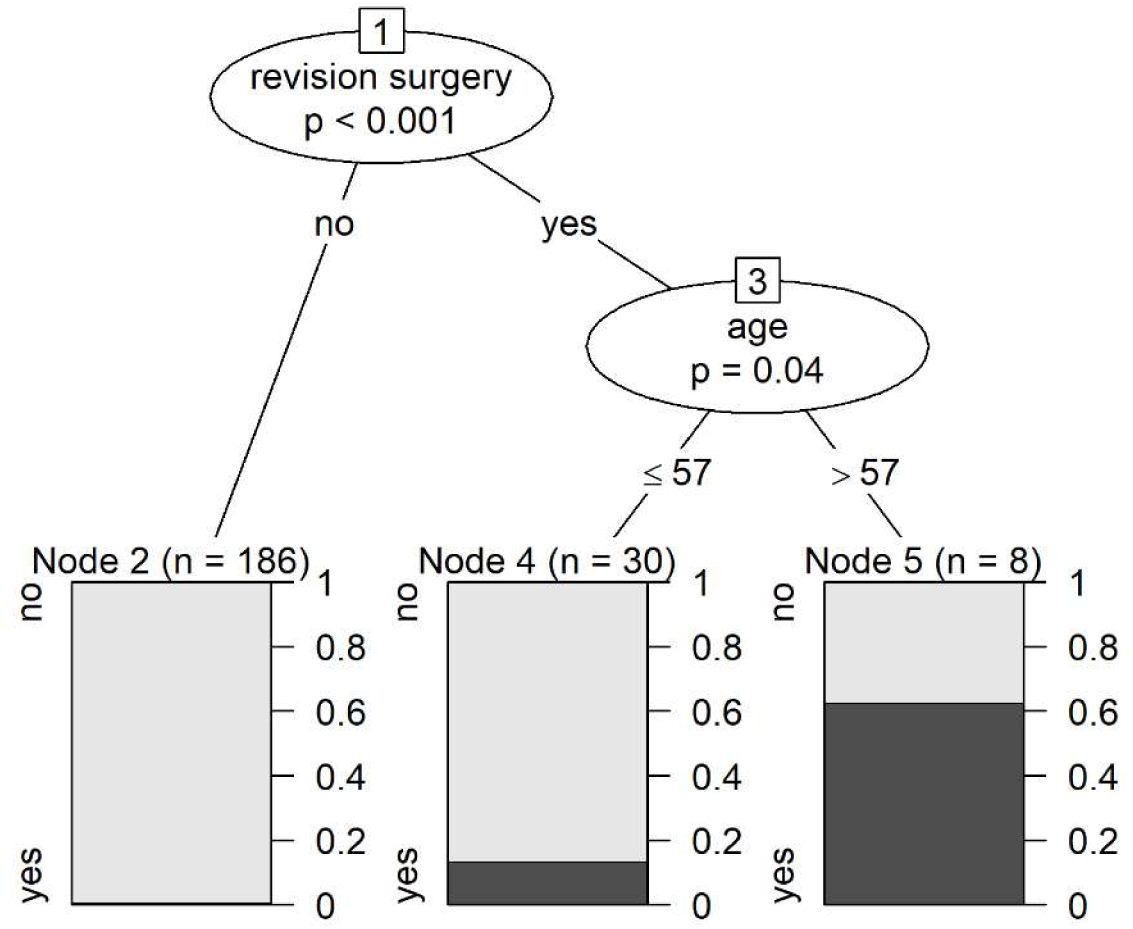
URP-CTREE first node: revision surgery (yes/no); third node: age at injury in years; three terminal nodes (2, 4, 5) for the endpoint SSI (yes/no); n, sample size; p, significance level; SSI, surgical site infection

#### SCIM

The functional outcome, assessed by the SCIM III, showed no significant change between the two observation periods. The mean (SD) change in SCIM between admission and discharge was 41.4(20.3) points in 2005-2009 and 42.0(19.9) in 2018-2022 (t-test p > 0.05). Also, the average SCIM at discharge was around 69 points in both observation periods (t-test p > 0.05; table 1). The change in SCIM between admission and discharge was significantly higher in individuals who received their initial spine surgery in the SCI center compared to those who underwent spine surgery in another hospital (mean (SD) 47.4 (21.6) vs. 38.9 (18.0); t-test p = 0.001).

The URP for the endpoint of SCIM at discharge identified two predictors for the functional outcome at hospital discharge: the AIS at admission and the number of levels fused. Individuals with an SCI classified as AIS D had the highest SCIM scores. Individuals with an SCI classified as AIS A, B, or C had a significantly higher SCIM (i.e. better functional outcome) when fusion length was two spinal segments or less compared to those where more than two segments have been fused (figure 3).

**Figure 3:**
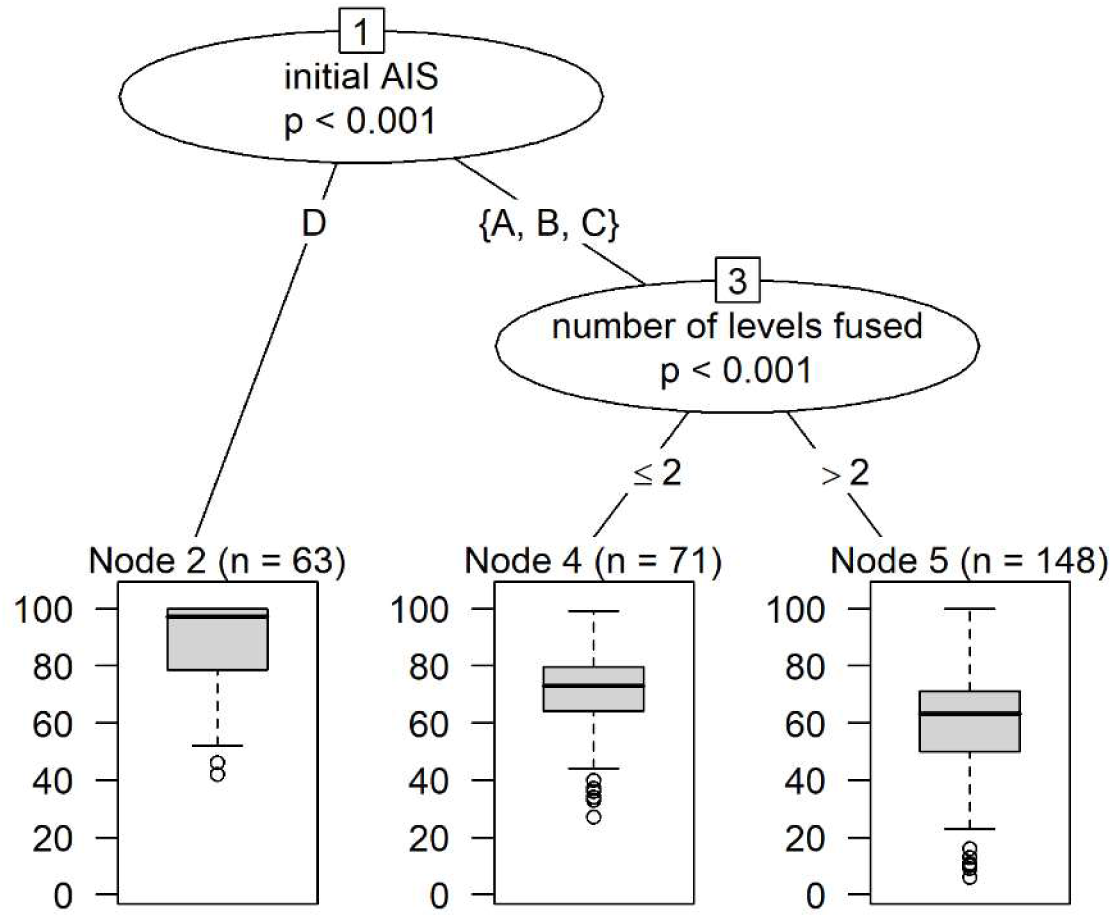
URP-CTREE first node: AIS at admission; third node: number of spinal levels fused; three terminal nodes (2, 4, 5) for the endpoint SCIM at discharge; n, sample size; p, significance level; AIS, American Spinal Injury Association (ASIA) Impairment Scale; SCIM, Spinal Cord Independence Measure

## Discussion

In our study cohort of 282 individuals with thoracic or thoracolumbar SCI, the overall rate of SSIs rose from 1.5% to 6.1% between the two observation periods (2005-2009 vs. 2018-2022). While this increase was not statistically significant, this trend, however, underscores a potential rise in infection risk associated with more extensive surgical procedures and more complex patient conditions in recent years. In our cohort, the number of spinal levels fused and the revision rate increased between the observation periods 2005-2009 and 2018-2022, reflecting potentially more complex injuries and/or changes in surgical practice. Also, both, the number of spinal segments fused and the revision rate were significantly associated with the occurrence of SSIs. While a biomechanically stable reconstruction of the anterior spinal column is particularly important in the thoracolumbar junction ^34^, these findings suggest that more extensive surgical interventions, characterized by longer fusion lengths, are associated with an increased risk of SSIs. This aligns with previous studies indicating that the extent of surgical intervention is a critical factor in the development of post-operative infections.^2,5–7,17,20,22^

When comparing patients who received their initial surgery at a specialized SCI center versus those treated initially at another hospital or trauma center, significant differences in fusion lengths were observed. Overall, patients treated in other hospitals had significantly longer fusions compared to those treated initially in a specialized SCI center. This difference was not observed in the earlier time period (2005-2009), but in the more recent period (2018-2022). Notably, fracture classification and the vertebral fracture level showed no significant differences between the centers, neither overall nor within the specific time periods (chi-squared p > 0.05). These findings imply that the type or level of injury did not account for the observed variations and surgical practices at other hospitals, including regional trauma centers, may have shifted toward more extensive spinal fusions.

Unbiased recursive partitioning by conditional inference (URP-CTREE) identified two primary predictors of SSIs: revision surgery and age. The need for revision surgery emerged as the strongest predictor, significantly increasing the risk of developing an SSI. Among patients who required revision surgery, those aged 57 years or older had an even higher likelihood of SSI, indicating that both surgical complexity and patient age are critical factors in increasing infection risk (figure 2). Additional patient-associated risk factors for SSIs are obesity, diabetes, and smoking.^6,9–16^ The risk of SSI increases by 6% with each added millimeter subcutaneous fat tissue between the laminal tissue of the vertebra and the skin ^13^, suggesting that obese patients are at a significantly increased risk.^13–16^ These findings highlight the importance of careful peri-operative management and possibly more aggressive prophylactic measures in older patients, obese patients, smokers, patients with diabetes, and those undergoing revision procedures.

### Functional outcome

The analysis of functional outcome, measured by the SCIM, revealed no significant changes between 2005-2009 and 2018-2022, with similar average SCIM scores at discharge across both observation periods. The URP-CTREE identified the initial severity of SCI (as classified by the AIS) and the number of spinal segments fused as significant predictors of functional outcome. Patients with less severe initial injuries (AIS D) had the best functional outcome. In those with more severe initial injuries (AIS A, B, or C), patients with shorter fusion lengths (two segments or less) achieved significantly better functional outcomes compared to those with longer fusions (figure 3). This suggests that, while extensive surgical interventions might be necessary in more complex cases, they may also be associated with reduced functional recovery.

While severe spinal instability or complex fractures constitute an indication for longer fusions, the data suggest that such extensive interventions may come at a cost. Longer fusion lengths are associated with a reduced range of motion in the spine, which can significantly limit a patient’s ability to perform activities of daily living and therefore their independence in daily life. The restriction in spinal mobility due to extensive fusions can lead to challenges in performing basic tasks such as bending, lifting, and even walking, thereby imposing limitations on both personal and social aspects of life. Previous studies have highlighted these concerns, indicating that the functional trade-offs associated with longer spinal fusions need careful consideration during surgical planning to balance stability with the preservation of post-operative quality of life.^35,36^

### Limitations

Previous literature has established that both diabetes and obesity are significant risk factors for the development of SSIs following spinal surgery. However, in this retrospective study, we were unable curate data on blood glucose levels, HbA1c, or body mass index. Future research should include these parameters to provide a more comprehensive understanding of these factors.

## Conclusion

In summary, revision surgery and patient age are significant predictors of SSIs in individuals with thoracic and thoracolumbar SCI. Additionally, the initial severity of SCI and the length of spinal fusion are important determinants of functional recovery, with shorter fusions being associated with better outcomes. The observed increase in fusion lengths and SSI rates over time warrants further investigation to optimize surgical approaches and improve patient outcomes.

## Ethics approval

All procedures performed in this study were in accordance with the Declaration of Helsinki and its later amendments, ICH GCP (International Conference on Harmonization Good Clinical Practice), the EU 2016/679 GDPR (General Data Protection Regulation), and the BDSG (Bundesdatenschutzgesetz) local legal regulations in Germany.

All individuals provided written informed consent permitting the use of their medical data for research purposes as part of their participation in the European Multicenter Study about Spinal Cord Injury (EMSCI) registry. The EMSCI has been approved by the Ethics Committee of the University of Heidelberg (Vote no. S-188/2003, Amendment 9).

## Data availability statement

The deidentified participant data utilized in this study, as well as additional related documents (study protocol, statistical analysis plan) can be made available upon reasonable request to the corresponding author. Access to the data will be subject to approval by institutional and ethical considerations.

## Author contribution statement

MV and IL conceived and designed the study. OM created a database for data curation and storage. MV and CW manually curated the data from various patient data management systems. OM, and IL have directly accessed and verified the underlying data. IL was responsible for statistical analysis. MK and IL were responsible for interpretation of results and writing of the manuscript. MV, OM, and DM performed critical revision of the manuscript regarding important intellectual content. All authors read and approved the final manuscript.

## Declaration of generative AI and AI-assisted technologies in the writing process

The authors acknowledge the use of large language models (ChaptGPT) to support the drafting of text within this article. Any suggestions obtained were critically reviewed and rephrased where needed. The authors emphasize that support was limited to grammatical and editorial phrasing. The authors take full responsibility for the content of the publication.

